## Supplementary Information for "Design of COVID-19 Staged Alert Systems to Ensure Healthcare Capacity with Minimal Closures"

Haoxiang Yang, Özge Sürer, Daniel Duque, David P. Morton, Bismark Singh, Spencer J. Fox, Remy Pasco, Kelly Pierce, Paul Rathouz, Zhanwei Du, Michael Pignone, Mark E. Escott, Stephen I. Adler, S. Claiborne Johnston, Lauren Ancel Meyers

#### A Material and Methods

Some cities in Texas, including Austin [1] and Houston [2], have staged systems to alert citizens of COVID-19 risks and inform mitigating behavior. Each stage has guidelines for the public, such as opening only essential businesses, avoiding gatherings of specified size, wearing face coverings, and avoiding dining and shopping. Stages with stricter distancing policies reduce disease transmission, but come with a higher socioeconomic cost. We optimize the timing of toggling between stages via triggers that monitor daily new hospital admissions while accounting for total hospitalizations and ICU hospitalizations. The model recommends a stricter stage of lock-down when the seven-day moving average of daily hospital admissions grows to exceed the stage's optimized *threshold*. A more relaxed stage is recommended when the same moving average drops below the stage's threshold. The sojourn time in each stage must be at least two weeks. We call this the "trigger policy."

To find the thresholds for each stage, we form a stochastic optimization model, in which disease dynamics are characterized by an enhanced SEIR-style model of disease transmission [3]. This epidemiological model comprises compartments for susceptible, exposed, pre-asymptomatic, pre-symptomatic, infectious-asymptomatic, infectious-symptomatic, infected and hospitalized in the general ward, infected and hospitalized in the intensive care unit (ICU), recovered, and deceased, which we denote by $S$, $E$, $PA$, $PY$, $IA$, $IY$, $IH$, $ICU$, $R$, and $D$, respectively. Fig. 1 diagrams the model's compartments and transitions for a metropolitan area. The population is partitioned into ten groups consisting of five age groups of low-risk and another five of high-risk individuals. Each group is represented with its own set of ten compartments (susceptible, exposed, etc.) so that in total the epidemiological model has 100 compartments.

We assume that the threshold for each stage is constant over time and is denoted by $\ell_i$ for stage $i$. The goal is to determine level $\ell_i$ so that the total expected cost of our trigger policy is minimized, while respecting epidemiological dynamics and hospital capacity using a probabilistic constraint. We approximate the expectation and probabilistic constraint by using Monte Carlo simulation to form a set of scenarios, indexed by $\omega$, and by using binary variables to count the number of paths that violate capacity and to determine the stage for each sample path at each point in time.

Notation:

*Indices and Sets*

| | |
|---|---|
| $t \in \mathcal{T}$ | set of time periods $\{1, 2, \ldots, \lvert\mathcal{T}\rvert\}$ [day] |
| $t \in \mathcal{T}_0$ | $\mathcal{T} \cup \{0\}$ |
| $a \in \mathcal{A}$ | set of age groups {0-4y, 5-17y, 18-49y, 50-64y, 65y+} |
| $r \in \mathcal{R}$ | risk groups $\{low, high\}$ |
| $i \in \mathcal{I}$ | predefined stages {1 (red), 2 (orange), 3 (yellow), 4 (blue)} |
| $\omega \in \Omega$ | set of scenarios |

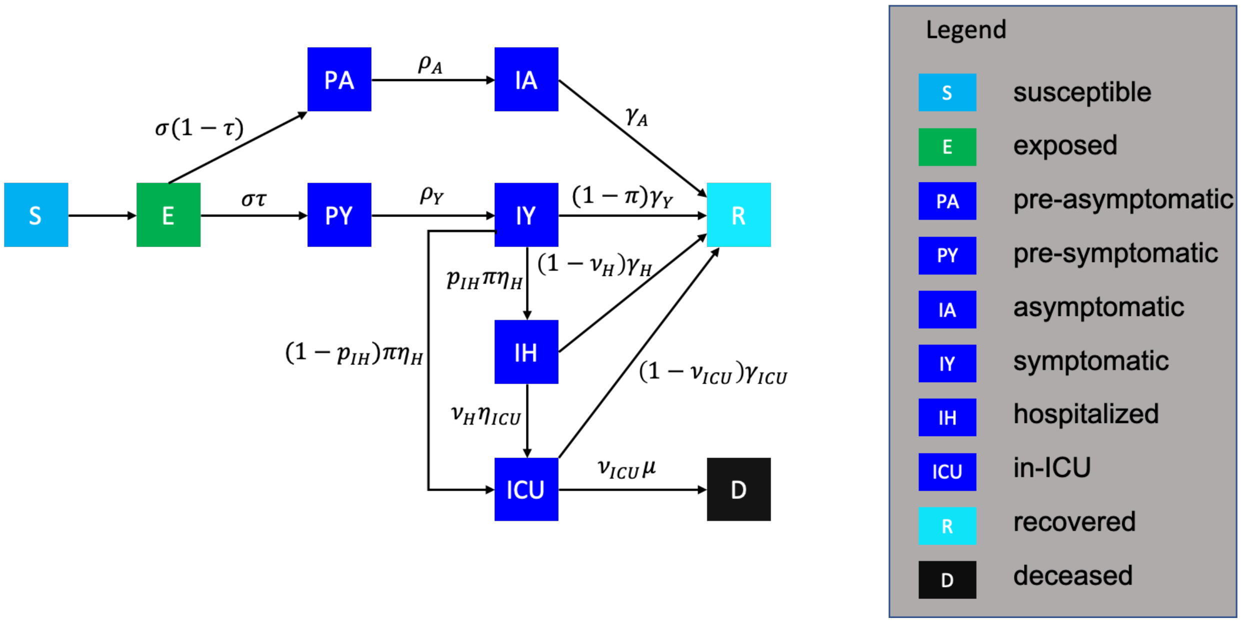


Supplementary Fig. 1: Diagram of the epidemiological model with compartments, transitions, and rates. The model has ten copies of such diagrams for each age-risk group pair, which interact to determine the (unmarked) rate of transition from compartment $S$ to $E$. Such transitions are governed by the transmission rate $\beta$ but also contact matrices and the current risk stage.

*Parameters*

*Epidemiological parameters*:

| | |
|---|---|
| $\beta$ | unmitigated transmission rate |
| $\sigma$ | rate at which exposed individuals become infectious |
| $\tau$ | proportion of exposed individuals who become symptomatic |
| $\rho_A$ | rate at which pre-asymptomatic individuals become asymptomatic |
| $\rho_Y$ | rate at which pre-symptomatic individuals become symptomatic |
| $\gamma_A$ | recovery rate from asymptomatic compartment |
| $\gamma_Y$ | recovery rate from symptomatic compartment |
| $\gamma_H^a$ | recovery rate from hospitalized compartment for age group $a$ |
| $\gamma_{ICU}^a$ | recovery rate from ICU compartment for age group $a$ |
| $P$ | proportion of pre-symptomatic transmission |
| $YHR^{a,r}$ | percent of symptomatic infectious that go to the hospital for age-risk group $a, r$ |
| $\eta_H$ | hospitalization rate after symptom onset |
| $\omega_A$ | infectiousness of individuals in $IA$ relative to $IY$ |
| $\omega_P^{a,r}$ | $\frac{P}{1-P}\frac{\tau(YHR^{a,r}/\eta_H+(1-YHR^{a,r})/\gamma_Y)+(1-\tau)\omega_A/\gamma_A}{\tau/\rho_Y+(1-\tau)\omega_A/\rho_A}$: infectiousness of pre-symptomatic individuals relative to $IY$ for age-risk group $a, r$ |
| $\pi^{a,r}$ | $\frac{\gamma_Y\cdot YHR^{a,r}}{[\eta_H-(\eta_H-\gamma_Y)YHR^{a,r}]}$: rate-adjusted proportion of symptomatic individuals who go to the hospital for age-risk group $a, r$ |
| $p_{IH}$ | percent of patients directly going to the general ward of the hospital |
| $HICUR$ | percent of general ward patients who get transferred to ICU |
| $\eta_{ICU}^a$ | ICU admission rate after admission to the general ward for age group $a$ |
| $\nu_H^a$ | $\frac{\gamma_H^a\cdot HICUR}{[\eta_{ICU}^a-(\eta_{ICU}^a-\gamma_H^a)HICUR]}$: rate-adjusted proportion of general ward patients transferred to ICU for age group $a$ |
| $\mu^a$ | rate from ICU to death for age group $a$ |
| $ICUFR^a$ | percent of hospitalized that die for age group $a$ |

$\nu^a_{ICU}$ $\frac{\gamma^a_{ICU} \cdot ICUFR^a}{[\mu^a - (\mu^a - \gamma^a_{ICU}) ICUFR^a]}$: ICU fatality rate-adjusted proportion for age group $a$

$\phi^{a',r',a,r}_{i,t}$ expected number of daily contacts from $(a', r')$ to $(a, r)$ at time $t$ under stage $i$

$N^{a,r}$ population of age-risk group $a, r$

*Additional parameters*:

$B$ number of hospital beds, including general ward and ICU, for COVID-19 patients

$B_{ICU}$ number of ICU beds for COVID-19 patients

$\varepsilon$ violation probability for probabilistic constraint

$C_i$ daily cost of being in stage $i$

*Variables*

*Epidemiological variables (for scenario $\omega \in \Omega$)*:

$S^{a,r}_{t,\omega}$ number of susceptible people of age group $a$, risk group $r$ at time $t$ [persons]

$dS^{a,r}_{t,\omega}$ $S^{a,r}_{t,\omega} - S^{a,r}_{t+1,\omega}$ [persons]

$E^{a,r}_{t,\omega}$ number of exposed people of age group $a$, risk group $r$ at time $t$ [persons]

$PA^{a,r}_{t,\omega}$ number of pre-asymptomatic people for $a, r, t$ [persons]

$PY^{a,r}_{t,\omega}$ number of pre-symptomatic people for $a, r, t$ [persons]

$IA^{a,r}_{t,\omega}$ number of infectious-asymptomatic people for $a, r, t$ [persons]

$IY^{a,r}_{t,\omega}$ number of infectious-symptomatic people for $a, r, t$ [persons]

$IH^{a,r}_{t,\omega}$ number of infected-hospitalized people in the general ward for $a, r, t$ [persons]

$ICU^{a,r}_{t,\omega}$ number of infected-hospitalized people in the ICU for $a, r, t$ [persons]

$R^{a,r}_{t,\omega}$ number of recovered people for $a, r, t$ [persons]

$D^{a,r}_{t,\omega}$ number of deceased people for $a, r, t$ [persons]

$H_{t,\omega}$ daily hospital admissions, from infectious-symptomatic to the general ward and ICU, at time $t$ [persons/day]

$\overline{H}_{t,\omega}$ seven-day moving average of $H_{t,\omega}$ [persons/day]

$U_{t,\omega}$ daily ICU admissions (from infectious-symptomatic and the general ward) at time $t$ [persons/day]

*Intervention variables*:

$\ell_i$ the stage-specific threshold (level) for the daily hospitalization rate

$X_{i,t,\omega}$ 1 if the system is in stage $i$ at time $t$ for scenario $\omega$; 0 otherwise

$Y_{i,t,\omega}$ 1 if the trigger statistic lies in range of stage $i$ at time $t$ for scenario $\omega$; 0 otherwise

$V_{i,t,\omega}$ 1 if $Y_{i,t,\omega} = 1$ prior to spending 14 days in the previous stage for scenario $\omega$; 0 otherwise

$Z_\omega$ 1 if healthcare capacity is exceeded in scenario $\omega$; 0 otherwise

We refer to Table 10 for further details on model parameters. We first define the epidemiological transition dynamics in the following equations for all $\omega \in \Omega$:

$$
\begin{aligned}
S^{a,r}_{t+1,\omega} - S^{a,r}_{t,\omega} =& - dS^{a,r}_{t,\omega} && \forall t \in \mathcal{T}_0, a \in \mathcal{A}, r \in \mathcal{R} && \text{[1a]}\\
E^{a,r}_{t+1,\omega} - E^{a,r}_{t,\omega} =& dS^{a,r}_{t,\omega} - \sigma E^{a,r}_{t,\omega} && \forall t \in \mathcal{T}_0, a \in \mathcal{A}, r \in \mathcal{R} && \text{[1b]}\\
PA^{a,r}_{t+1,\omega} - PA^{a,r}_{t,\omega} =& \sigma(1-\tau)E^{a,r}_{t,\omega} - \rho_A PA^{a,r}_{t,\omega} && \forall t \in \mathcal{T}_0, a \in \mathcal{A}, r \in \mathcal{R} && \text{[1c]}\\
IA^{a,r}_{t+1,\omega} - IA^{a,r}_{t,\omega} =& \rho_A PA^{a,r}_{t,\omega} - \gamma_A IA^{a,r}_{t,\omega} && \forall t \in \mathcal{T}_0, a \in \mathcal{A}, r \in \mathcal{R} && \text{[1d]}\\
PY^{a,r}_{t+1,\omega} - PY^{a,r}_{t,\omega} =& \sigma\tau E^{a,r}_{t,\omega} - \rho_Y PY^{a,r}_{t,\omega} && \forall t \in \mathcal{T}_0, a \in \mathcal{A}, r \in \mathcal{R} && \text{[1e]}\\
IY^{a,r}_{t+1,\omega} - IY^{a,r}_{t,\omega} =& \rho_Y PY^{a,r}_{t,\omega} - (1-\pi^{a,r})\gamma_Y IY^{a,r}_{t,\omega} - \pi^{a,r}\eta_H IY^{a,r}_{t,\omega} && \forall t \in \mathcal{T}_0, a \in \mathcal{A}, r \in \mathcal{R} && \text{[1f]}\\
IH^{a,r}_{t+1,\omega} - IH^{a,r}_{t,\omega} =& p_{IH}\pi^{a,r}\eta_H IY^{a,r}_{t,\omega} - (1-\nu^a_H)\gamma^a_H IH^{a,r}_{t,\omega} - \nu^a_H \eta^a_{ICU} IH^{a,r}_{t,\omega} && \forall t \in \mathcal{T}_0, a \in \mathcal{A}, r \in \mathcal{R} && \text{[1g]}\\
ICU^{a,r}_{t+1,\omega} - ICU^{a,r}_{t,\omega} =& (1-p_{IH})\pi^{a,r}\eta_H IY^{a,r}_{t,\omega} + \nu^a_H \eta^a_{ICU} IH^{a,r}_{t,\omega} - && && \text{[1h]}\\
& (1-\nu^a_{ICU})\gamma^a_{ICU} ICU^{a,r}_{t,\omega} - \nu^a_{ICU}\mu^a ICU^{a,r}_{t,\omega} && \forall t \in \mathcal{T}_0, a \in \mathcal{A}, r \in \mathcal{R} && \text{[1i]}\\
R^{a,r}_{t+1,\omega} - R^{a,r}_{t,\omega} =& \gamma_A IA^{a,r}_{t,\omega} + (1-\pi^{a,r})\gamma_Y IY^{a,r}_{t,\omega} + (1-\nu^a_H)\gamma^a_H IH^{a,r}_{t,\omega} + && && \text{[1j]}\\
& (1-\nu^a_{ICU})\gamma^a_{ICU} ICU^{a,r}_{t,\omega} && \forall t \in \mathcal{T}_0, a \in \mathcal{A}, r \in \mathcal{R} && \text{[1k]}\\
D^{a,r}_{t+1,\omega} - D^{a,r}_{t,\omega} =& \nu^a_{ICU}\mu^a ICU^{a,r}_{t,\omega} && \forall t \in \mathcal{T}_0, a \in \mathcal{A}, r \in \mathcal{R} && \text{[1l]}\\
dS^{a,r}_{t,\omega} =& S^{a,r}_{t,\omega} \sum_{a' \in \mathcal{A}} \sum_{r' \in \mathcal{R}} \sum_{i \in \mathcal{I}} \frac{\beta \phi^{a',r',a,r}_{i,t} X_{i,t,\omega}}{N^{a',r'}} \Big( IY^{a',r'}_t + \omega_A IA^{a',r'}_t + && && \\
& \omega^{a',r'}_P \omega_A PA^{a',r'}_t + \omega^{a',r'}_P PY^{a',r'}_t \Big) && \forall t \in \mathcal{T}_0, a \in \mathcal{A}, r \in \mathcal{R}. && \text{[1m]}
\end{aligned}
$$

The initial conditions, for analysis in both Austin and Houston, have all variables indexed by $t = 0$ as zero except the following:

$$IY^{18\text{-}49,low}_{0,\omega} = 1, S^{18\text{-}49,low}_{0,\omega} = N^{18\text{-}49,low} - 1, \text{and } S^{a,r}_{0,\omega} = N_{a,r} \forall (a,r) \in \mathcal{A} \times \mathcal{R} \setminus \{(18\text{-}49, low)\}. \qquad [2]$$

The epidemiological dynamics largely follow the formulation used in [4] with the addition of three compartments to improve model fidelity and to distinguish beds in the ICU and general ward. The initial conditions specify a single infectious individual in the 18-49 age group with low risk. The age-risk groups are initialized with the rest of the population in their respective susceptible compartments. Eqs. [1a]-[1m] then provide a sample path, indexed by $\omega$, for the progression of the disease in the community. For the moment, the indicator variables $X_{i,t,\omega} \in \{0, 1\}$ are taken as input, and select the current stage and, in turn, the expected number of daily contacts via $\phi^{a',r',a,r}_{i,t}$. The contact matrices are indexed by $t$ because they capture whether school is currently open and if so, the school calendar; they further capture weekdays versus weekends and the level of cocooning, which can vary with time; and they capture contacts at school, home, work, and another catch-all category. We assume that sufficient precautions are taken in hospitals so that hospitalized cases do *not* contribute to infecting others via Eq. [1m]. The most significant updates of the model from that in [4] are in additional compartments. We use constructs similar to He et al. [5] for a pre-symptomatic period to more accurately model the profile of infectiousness of individuals by including pre-symptom onset transmission. We also model the ICU compartment explicitly for two reasons. First, patients in the ICU have different durations in the hospital than those in the general ward, and second it allows us to account for ICU capacity as a resource. We let $p_{IH}$ denote the probability a hospitalized patient is admitted to a general ward bed and the remaining fraction go directly to the ICU. As Fig. 1 and Eq. [1h] indicate, it is possible to transfer general ward patients to the ICU later if needed. All deaths are recorded from the ICU.

The optimization model can be formulated as follows:

$$
\begin{aligned}
\min \quad & \frac{1}{|\Omega|}\sum_{\omega\in\Omega}\sum_{i\in\mathcal{I}}\sum_{t\in\mathcal{T}} C_i X_{i,t,\omega} && && \text{[3a]}\\
\text{s.t.} \quad & \text{Equations [1] and [2]} && && \text{[3b]}\\
& H_{t,\omega} = \sum_{a\in\mathcal{A}}\sum_{r\in\mathcal{R}} \pi^{a,r}\eta_H IY^{a,r}_{t,\omega} && \forall t\in\mathcal{T},\omega\in\Omega && \text{[3c]}\\
& \overline{H}_{t,\omega} = \frac{1}{7}\sum_{t'=\max\{t-6,1\}}^{t} H_{t,\omega} && \forall t\in\mathcal{T},\omega\in\Omega && \text{[3d]}\\
& \ell_i - M(1-Y_{i,t,\omega}) \le \overline{H}_{t,\omega} \le \ell_{i+1} + M(1-Y_{i,t,\omega}) && \forall i\in\mathcal{I}, t\in\mathcal{T},\omega\in\Omega && \text{[3e]}\\
& 2V_{i,t,\omega} - 1 \le X_{i,t-1,\omega} - X_{i,\max\{t-14,0\},\omega} \le V_{i,t,\omega} && \forall i\in\mathcal{I}, t\in\mathcal{T},\omega\in\Omega && \text{[3f]}\\
& X_{i,t,\omega} \ge V_{i,t,\omega} && \forall i\in\mathcal{I}, t\in\mathcal{T},\omega\in\Omega && \text{[3g]}\\
& X_{i,t,\omega} \le Y_{i,t,\omega} + V_{i,t,\omega} && \forall i\in\mathcal{I}, t\in\mathcal{T},\omega\in\Omega && \text{[3h]}\\
& \sum_{i\in\mathcal{I}} X_{i,t,\omega} = 1 && \forall t\in\mathcal{T},\omega\in\Omega && \text{[3i]}\\
& X_{i,0,\omega} = 0 && \forall i\in\mathcal{I},\omega\in\Omega && \text{[3j]}\\
& \sum_{a\in\mathcal{A}}\sum_{r\in\mathcal{R}} ICU^{a,r}_{t,\omega} \le B_{ICU} + MZ_\omega && \forall t\in\mathcal{T},\omega\in\Omega && \text{[3k]}\\
& \sum_{\omega\in\Omega} Z_\omega \le \lfloor \varepsilon|\Omega| \rfloor && && \text{[3l]}\\
& 0 \le \ell_i \le \ell_{i-1} && \forall i\in\mathcal{I}\setminus\{1\} && \text{[3m]}\\
& X_{i,t,\omega}, V_{i,t,\omega}, Y_{i,t,\omega} \in \{0,1\} && \forall i\in\mathcal{I}, t\in\mathcal{T},\omega\in\Omega && \text{[3n]}\\
& Z_\omega \in \{0,1\} && \forall \omega\in\Omega. && \text{[3o]}
\end{aligned}
$$

For simplicity, we write the finite-difference Eqs. [1] in a deterministic form. They become stochastic, and require indexing by $\omega$, because binomial random variables replace terms like $\sigma E^{a,r}_{t,\omega}$; here the binomial random variable has parameter $n = E^{a,r}_{t,\omega}$ and $\sigma$ serves as the "success" probability. This construct is pervasive throughout right-hand side terms in Eqs. [1]. In addition to these "micro" stochastics there are "macro" stochastics because we model $\sigma$, $\omega_A$, $\gamma_A$, and $\gamma_Y$ as random variables that are subject to a Monte Carlo draw at time 0 of the simulation. We discuss this further in Appendix C.

For each sample path, $\omega$, we record the number of daily hospital admissions and its seven-day moving average in constraints [3c] and [3d], aggregated across the general ward and ICU. At time $t$, the stage is decided using two criteria. We are nominally in stage $i$ if the trigger statistic lies between the threshold of stage $i$, $\ell_i$, that of stage $i+1$, $\ell_{i+1}$. This occurs unless we would spend fewer than 14 days in our current stage, in which case we cannot yet switch stages. This logic is enforced by constraints [3e]-[3j]. Constraint [3e] identifies the would-be stage based on the trigger statistic, which is the seven-day moving average of the hospital admissions. The big-$M$ coefficient in constraint [3e] stands for a sufficiently large number. Constraint [3f] forces variable $V_{i,t,\omega}$ to take value 1 if we switched to stage $i$ in the previous 14 days and value 0 otherwise, and thus it forces the system to stay in the same stage through constraint [3g]. If $V_{i,t,\omega} = 0$ then constraint [3h] means that the stage is decided by the trigger statistic through variable $Y_{i,t,\omega}$. Constraint [3i] states that only one stage can be selected at any time-scenario ($t$,$\omega$) pair.

The expected cost is calculated in the objective function [3a] by summing the daily socioeconomic costs over the time horizon. Constraint [3k] identifies scenarios for which, at some time point, the number of ICU patients exceeds the ICU capacity, where $M$ is again sufficiently large so the constraint is vacuous if $Z_\omega = 1$. Constraint [3l] ensures the probability such a violation occurs is at most fraction $\varepsilon$ of the total scenarios. Here we use the number of ICU patients in the probabilistic constraint, but it can be replaced by the number of total hospitalized patients, respectively replacing $ICU^{a,r}_{t,\omega}$ and $B_{ICU}$ by $ICU^{a,r}_{t,\omega} + IH^{a,r}_{t,\omega}$ and $B$.

We have formulated the model with daily time periods, which simplifies notation for computing the seven-day moving average of new admissions, the logic behind the rule of spending two weeks in a stage, etc. That said, in implementation we use ten time steps per day, which suffices for the fidelity of the epidemiological dynamics in Eqs. [1].

We generate 300 scenarios for $\Omega$ according to the procedure described in Appendix C. After obtaining a set of optimal triggers, we generate another 300 scenarios, $\Omega'$, to assess performance. Those two sets of scenarios are similar to "training" and "testing" data used in statistics and machine learning. As a result, it is possible to obtain thresholds that meet the probabilistic constraint under $\Omega$, but violate that constraint when tested using $\Omega'$, although in our experience such violations are both rare and modest.

Model [3] is a large-scale stochastic mixed-integer nonlinear program. Problems of the scale we consider cannot currently be solved using commercial integer programming software. We approximately solve the model using a grid search procedure. For a fixed set of thresholds, $\ell_i, i \in \mathcal{I}$, which satisfy inequalities [3m], we can run the simulation model for all $\omega \in \Omega$ in parallel, applying transition dynamics [1]-[2] and using the logic of [3c]-[3k] and [3n]-[3o] to compute the binary variables which indicate the stages, $X_{i,t,\omega}$, and the scenarios with capacity violations, $Z_\omega$. For each set of thresholds that we consider in our grid search, we select the configuration that yields the minimum expected cost [3a] while satisfying the probabilistic constraint [3l]. Our grid search considers all configurations of the thresholds $\ell_i, i \in \mathcal{I}$, using increments of 10 that satisfy [3m] after computing an upper bound on $\ell_i$ necessary to satisfy the probabilistic constraint [3l].

### B Model Parameters

Tables 1 and 2 partition the population of the Austin and Houston MSAs based on age groups (0-4 years old, 5-17 years old, 18-49 years old, 50-64 years old, and 65 years and older) and risk groups (low risk and high risk). The high-risk group proportions are estimated based on the population with chronic conditions listed by the CDC 500 cities data [6]. Population data processing is detailed in the appendix of [4] and here we present only the final numbers used for this paper's analysis.

| $N^{a,r}$ | 0-4 | 5-17 | 18-49 | 50-64 | 65 and older |
|---|---|---|---|---|---|
| Low risk | 128527 | 327148 | 915894 | 249273 | 132505 |
| High risk | 9350 | 37451 | 156209 | 108196 | 103763 |

Supplementary Table 1: Austin age-risk group populations.

| $N^{a,r}$ | 0-4 | 5-17 | 18-49 | 50-64 | 65 and older |
|---|---|---|---|---|---|
| Low risk | 503894 | 1101232 | 2701594 | 778587 | 373036 |
| High risk | 38695 | 136609 | 624121 | 395585 | 344031 |

Supplementary Table 2: Houston age-risk group populations.

We define four baseline contact matrices, $\mathcal{H}, \mathcal{S}, \mathcal{W}$, and $\mathcal{O}$, to describe the contact frequency between age groups at home, at school, at work, and at other locations. These *baseline* matrices assume there is no difference in contacts among the low- and high-risk groups. Each row and column represents an age group, in the order of 0-4 years old, 5-17 years old, 18-49 years old, 50-64 years old, and 65 years old and above, with the row-column value corresponding to a "from-to" transmission contact:

$$\mathcal{H} = \begin{bmatrix} 0.5 & 0.9 & 2.0 & 0.1 & 0.0 \\ 0.2 & 1.7 & 1.9 & 0.2 & 0.0 \\ 0.2 & 0.9 & 1.7 & 0.2 & 0.0 \\ 0.2 & 0.7 & 1.2 & 1.0 & 0.1 \\ 0.1 & 0.7 & 1.0 & 0.3 & 0.6 \end{bmatrix} \qquad \mathcal{S} = \begin{bmatrix} 1.0 & 0.5 & 0.4 & 0.1 & 0.0 \\ 0.2 & 3.7 & 0.9 & 0.1 & 0.0 \\ 0.0 & 0.7 & 0.8 & 0.0 & 0.0 \\ 0.1 & 0.8 & 0.5 & 0.1 & 0.0 \\ 0.0 & 0.0 & 0.1 & 0.0 & 0.0 \end{bmatrix}$$

$$\mathcal{W} = \begin{bmatrix} 0.0 & 0.0 & 0.0 & 0.0 & 0.0 \\ 0.0 & 0.1 & 0.4 & 0.0 & 0.0 \\ 0.0 & 0.2 & 4.5 & 0.8 & 0.0 \\ 0.0 & 0.1 & 2.8 & 0.9 & 0.0 \\ 0.0 & 0.0 & 0.1 & 0.0 & 0.0 \end{bmatrix} \qquad \mathcal{O} = \begin{bmatrix} 0.7 & 0.7 & 1.8 & 0.6 & 0.3 \\ 0.2 & 2.6 & 2.1 & 0.4 & 0.2 \\ 0.1 & 0.7 & 3.3 & 0.6 & 0.2 \\ 0.1 & 0.3 & 2.2 & 1.1 & 0.4 \\ 0.0 & 0.2 & 1.3 & 0.8 & 0.6 \end{bmatrix}.$$

The contact matrices $\phi_{i,t}^{a',r',a,r}$ are calculated in the same way as Table S6 in [4], considering the effect of weekends, holidays, school closures, and physical distancing and cocooning of high-risk populations based on the risk stage. Stages correspond to distancing stages of different strictness, which govern the reduced number of daily contacts people make relative to baseline. In our model, this is reflected by a coefficient $\kappa_i, i \in \mathcal{I}$, where $\kappa_i = 0.75$ would reduce the expected number of contacts to 25% of the baseline value. For the age group of 65 years and older and for the high-risk group, we use reductions based on cocooning, which are represented by coefficients $c_i, i \in \mathcal{I}$:

$$\phi_{i,t}^{a',r',a,r} = \begin{cases} (1-\kappa_i)\left[(1-1_{\{\text{off day}\}})\cdot(1-1_{\{\text{school closure}\}})\cdot\mathcal{S}_{a',a}+ \right. & \text{if } a',a \in \{\text{0-4yr, 5-17yr, 18-49yr, 50-64yr}\}, \\ \quad \left. (1-1_{\{\text{off day}\}})\cdot\mathcal{W}_{a',a}+\mathcal{H}_{a',a}+\mathcal{O}_{a',a}\right] & \quad r',r \neq \text{high-risk} \\ (1-c_i)\left[(1-1_{\{\text{off day}\}})\cdot(1-1_{\{\text{school closure}\}})\cdot\mathcal{S}_{a',a}+ \right. & \\ \quad \left. (1-1_{\{\text{off day}\}})\cdot\mathcal{W}_{a',a}+\mathcal{H}_{a',a}+\mathcal{O}_{a',a}\right] & \text{otherwise.} \end{cases} \qquad [4]$$

The indicator $1_{\{\text{off day}\}}$ takes value 1 if the day is a weekend or holiday and is otherwise 0, and a similar indicator accounts for school closures. When a high-risk group, along with those 65 years and older, is involved either on the "giving" or "receiving" end of a contact, Eq. [4] assumes reduced transmission via the cocooning coefficient, $c_i$.

The following are key dates during the pandemic in Texas, and some define time blocks, which we use in estimating time-varying transmission reduction factors and other key model parameters as we describe shortly:

- February 19, 2020: Seed date for simulation of Houston, assuming seeding by a single symptomatic individual age 18-49y. This corresponds to 14 days prior to the first detected COVID-19 case in Houston on March 4, 2020.
- February 28, 2020: Seed date for simulation of Austin, assuming seeding by a single symptomatic individual age 18-49y. This corresponds to 14 days prior to the first detected COVID-19 case in Austin on March 13, 2020.
- March 24, 2020: Austin's Stay Home-Work Safe Order is enacted at midnight [7]. On the same day, Harris County (Houston) issues Stay Home, Work Safe Order [8].
- May 1, 2020: The Governor of Texas relaxed physical distancing orders statewide [9].
- May 21, 2020: Just prior to Memorial Day Weekend.
- June 26, 2020: The Governor of Texas issued an executive order limiting service at bars and restaurants, and Travis County (which includes Austin) banned gatherings of more than 100 people [10, 11].
- July 17, 2020: Time point in hospitalization data suggesting a change in dynamics.
- August 9, 2020: The last day of observed data from the hospital system used in estimating changes in ICU dynamics.
- August 20, 2020: First day students returned to residence halls at the University of Texas at Austin.

- October 7, 2020: The last day of observed data used in estimating model parameters.

We assume that there are six time blocks denoted by $\mathcal{T}_j$ for $j \in \{1, 2, 3, 4, 5, 6\}$ as defined in Table 3. They guide fitting of transmission-reduction parameters, $\kappa$ and $c$, and certain dynamics in use of the ICU and hospital duration, as detailed below.

| Time Block | Start Date | End Date | Definition |
|---|---|---|---|
| $\mathcal{T}_1$ | 2/28/20 (Austin) | 3/23/20 | unmitigated transmission before first stay-home order |
| | 2/19/20 (Houston) | 3/23/20 | |
| $\mathcal{T}_2$ | 3/24/20 | 5/20/20 | effective period for first stay-home order |
| $\mathcal{T}_3$ | 5/21/20 | 6/25/20 | relaxed period starting with Memorial Day weekend |
| $\mathcal{T}_4$ | 6/26/20 | 7/16/20 | period of effective physical distancing |
| $\mathcal{T}_5$ | 7/17/20 | 8/19/20 | period distinguished by changes in ICU dynamics |
| $\mathcal{T}_6$ | 8/20/20 | 10/7/20 | period of effective physical distancing |

Supplementary Table 3: The five time blocks, $\mathcal{T}_1$, $\mathcal{T}_2$, $\mathcal{T}_3$, $\mathcal{T}_4 \cup \mathcal{T}_5$, and $\mathcal{T}_6$ correspond to different rates of spread, as estimated using transmission-reduction factors $\kappa$ and $c$. The fourth and fifth time blocks, $\mathcal{T}_4$ and $\mathcal{T}_5$, differ only in dynamics involving the ICU, both the admission probability and the sojourn time in the general ward prior to ICU admission.

We model the hospitalization dynamics, including proportions of hospitalized requiring the ICU and durations in the general ward and ICU, using data from a multi-facility hospital system serving the central Texas region, including Austin, Texas ("hospital system data"). While we model differences based on five age groups, we assume the same hospital dynamics in different hospital systems after a patient is admitted across Austin and Houston due to similar medical standards. Conditional on being admitted to the hospital, we observe a decreasing trend in the probability a patient is admitted to the ICU throughout the time horizon, which holds for both direct admissions to the ICU and patients who are first admitted to the general ward. Among patients who enter the general ward and are then admitted to the ICU, their duration of stay in the general ward, determined by $\eta_{ICU}$, grows over time. For each time block, $\mathcal{T}_j$, we assume a constant $\eta_{ICU,j}$ and further assume a constant daily decrease, $r_j$, on both of the fractions, $p_{IH}$ and $HICUR$:

$$p_{IH,t+1} = r_j p_{IH,t} \qquad \forall j \in \{1, 2, 3, 4, 5, 6\}, t \in \mathcal{T}_j \qquad [5a]$$

$$HICUR_{t+1} = r_j HICUR_t \qquad \forall j \in \{1, 2, 3, 4, 5, 6\}, t \in \mathcal{T}_j, \qquad [5b]$$

along with a similar decrement across boundaries of the blocks. We use duration times for each time block from the hospital system data to estimate $\eta^a_{ICU,j}$ and fit $r_j$, with the estimated parameters in Table 4.

| | age group | $\mathcal{T}_1$ | $\mathcal{T}_2$ | $\mathcal{T}_3$ | $\mathcal{T}_4$ | $\mathcal{T}_5 \cup \mathcal{T}_6$ |
|---|---|---|---|---|---|---|
| $\eta^a_{ICU,j}$ | 0-4 yr | 0.5882 | 0.5882 | 0.3885 | 0.2640 | 0.2589 |
| | 5-17 yr | 0.5882 | 0.5882 | 0.3885 | 0.2640 | 0.2589 |
| | 18-49 yr | 0.5882 | 0.5882 | 0.3885 | 0.2640 | 0.2589 |
| | 50-64 yr | 0.6273 | 0.6273 | 0.4143 | 0.2815 | 0.2761 |
| | $\geq 65$ yr | 0.6478 | 0.6478 | 0.4278 | 0.2907 | 0.2851 |
| $r_j$ | | 0.9973 | 0.9973 | 0.9932 | 0.9921 | 1 |

Supplementary Table 4: Estimates of ICU admission probability parameters, $\eta_{ICU}, p_{IH}$, and $HICUR$; see Fig. 1 and accompanying parameter definitions. For each age group, $a$, and each time block, $j$, we specify $\eta_{ICU}$, and we give the daily decrement factor, $r_j$, used in Eq. [5].

Using the hospital system data, and consistent with the transition diagram in Fig. 1, we define the ICU duration for a patient as the time between their admission to the ICU and their discharge from the hospital. The reality is more complex as ICU patients typically return to the general ward prior to discharge from the hospital, and iterations between the two units, driven by a patient's health status, can also occur. Therefore, the reported duration in the ICU leads to over estimating ICU utilization and under-estimating that of the general ward. To handle this in our model, we introduce two constant parameters, $\alpha_{ICU}$ and $\alpha_H$, to better estimate durations in the ICU and general ward and better represent their respective utilization:

$$\gamma_H = (1 - \alpha_H)\gamma_H^0$$
$$\gamma_{ICU} = (1 + \alpha_{ICU})\gamma_{ICU}^0$$
$$\mu = (1 + \alpha_{ICU})\mu^0,$$

where $\gamma_H^0$, $\gamma_{ICU}^0$, and $\mu^0$ are obtained from the hospital system data, with each row corresponding to an age group in ascending order:

$$\gamma_H^0 = \begin{bmatrix} 0.2399 \\ 0.2399 \\ 0.2399 \\ 0.2222 \\ 0.2124 \end{bmatrix}, \ \gamma_{ICU}^0 = \begin{bmatrix} 0.0700 \\ 0.0700 \\ 0.0700 \\ 0.0575 \\ 0.0518 \end{bmatrix}, \ \mu^0 = \begin{bmatrix} 0.0749 \\ 0.0749 \\ 0.0749 \\ 0.0766 \\ 0.0799 \end{bmatrix},$$

with units of day$^{-1}$.

The bulk of the epidemiological and hospitalization parameters are specified above or are detailed in Tables 10 and 11, with the latter obtained from the literature or information collected from local healthcare agencies. The time blocks are specified in Table 3. Given these, we estimate 14 parameters, but with 7 degrees of freedom, as we detail below. We perform the fit of the deterministic SEIR model in Eqs. [1] using: (i) daily COVID-19 admissions, denoted $H_t$ and available for Austin only; (ii) a daily COVID census in the general ward, $IH_t$; and (iii) a daily COVID census in the ICU, $ICU_t$, all on day $t$. The fit is performed separately for the Austin MSA and Houston MSA. By minimizing a weighted sum of least-square errors, we estimate $\hat{\kappa}_j$ and $\hat{c}_j$, $j = 1, 2, \ldots, 6$, $\alpha_H$, and $\alpha_{ICU}$, using SciPy/Python [12] via `scipy.optimize.least_squares`. We use the "hat" notation on $\kappa$ and $c$ to distinguish, for example, $\hat{\kappa}_j$ for *time block* $j$ (see Table 3) from $\kappa_i$, which corresponds to *stage* $i$ per Eq. [4], and show the mapping shortly.

We minimize

$$\sum_t (IH_t - \widehat{IH}_t)^2 + w_{ICU}^2 \sum_t (ICU_t - \widehat{ICU}_t)^2 + w_H^2 \sum_t (H_t - \widehat{H}_t)^2,$$

where $\widehat{IH}_t$, $\widehat{ICU}_t$, and $\widehat{H}_t$ denote the estimated $IH_t$, $ICU_t$, and $H_t$ obtained through Eqs. [1]; $w_{ICU}$ and $w_H$ are scaling constants; and the sum is over $t \in \mathcal{T}_1 \cup \cdots \cup \mathcal{T}_6$. We assume $w_{ICU} = 1.50$ and $w_H = 7.58$, as those values approximate magnitudes relative to that of the general ward. To obtain a parsimonious model, we use $\hat{c}_1 = 0$, $\hat{c}_2 = \hat{c}_3 = \hat{\kappa}_2$, $\hat{c}_4 = \hat{c}_5 = \hat{\kappa}_4 = \hat{\kappa}_5$ and $\hat{c}_6 = \hat{\kappa}_6$, which reduces the number of estimated parameters from 14 to 7. The rationale is that there was effectively no cocooning during the initial time block $\mathcal{T}_1$, and thus we set $\hat{c}_1 = 0$. Because physical distancing was stricter over time block $\mathcal{T}_2$ than time block $\mathcal{T}_3$, and the cocooning effectiveness parameters are expected to be at least that of the distancing parameters, we use $\hat{c}_2 = \hat{c}_3 = \hat{\kappa}_2$. Because of behavioral changes over time, including increased use of face-masks, when we perform the fit we observe greater reduction in transmission in time blocks 4 and 5 than we do in block 2, and so we use $\hat{c}_4 = \hat{\kappa}_4$. Finally, while hospitalization parameters differ over time blocks $\mathcal{T}_4$ and $\mathcal{T}_5$, rates of transmission do not appear to differ significantly, and so we consider $\hat{c}_4 = \hat{c}_5 = \hat{\kappa}_4 = \hat{\kappa}_5$.

We use the trust region reflective algorithm (`trf`) in `scipy.optimize.least_squares`, with lower and upper bounds on each parameter of 0 and 1, respectively. The algorithm obtains locally optimal values of the parameters, the quality of which has been validated by comparing projections with the observed data. All the remaining parameters are set to their default values (see above and Tables 10 and 11). The fitted values for $\hat{\kappa}_j$ and $\hat{c}_j$ and $\alpha_H$ and $\alpha_{ICU}$ are given in Table 5 for both Austin and Houston.

The physical distancing parameters for each stage are mapped to $\kappa_i$ and $c_i$ for $i \in \mathcal{I}$ based on the historical

| | Austin | | Houston | |
|---|---|---|---|---|
| $j$ | $\hat{\kappa}_j$ | $\hat{c}_j$ | $\hat{\kappa}_j$ | $\hat{c}_j$ |
| 1 | 0.0613 | 0.0000 | 0.1168 | 0.0000 |
| 2 | 0.7436 | 0.7436 | 0.7304 | 0.7304 |
| 3 | 0.6026 | 0.7436 | 0.5643 | 0.7304 |
| 4 | 0.7815 | 0.7815 | 0.7553 | 0.7553 |
| 5 | 0.7815 | 0.7815 | 0.7553 | 0.7553 |
| 6 | 0.7544 | 0.7544 | 0.7118 | 0.7118 |
| $\alpha_H$ | 0.2862 | | 0.4858 | |
| $\alpha_{ICU}$ | 0.5621 | | 0.4876 | |

Supplementary Table 5: Fitted transmission reduction parameters, $\hat{\kappa}_j$, and cocooning effectiveness parameters, $\hat{c}_j$, for each time block $\mathcal{T}_j$, along with estimated hospitalization duration adjustment parameters, $\alpha_H$ and $\alpha_{ICU}$, for both Austin and Houston.

implementation of the policy. We set:

$$\kappa_1 = \hat{\kappa}_4 \qquad c_1 = \hat{\kappa}_4 \tag{6a}$$

$$\kappa_2 = \frac{\hat{\kappa}_4 + \hat{\kappa}_3}{2} \qquad c_2 = \hat{\kappa}_2 \tag{6b}$$

$$\kappa_3 = \hat{\kappa}_3 \qquad c_3 = \hat{\kappa}_2 \tag{6c}$$

$$\kappa_4 = \hat{\kappa}_3 - \frac{\hat{\kappa}_4 - \hat{\kappa}_3}{2} \qquad c_4 = 1 - 1.25 \cdot (1 - c_2). \tag{6d}$$

In Eq. [6a] we set $\kappa_1$ (red) to the strictest observed level of transmission reduction, June 26–August 19, 2020. In Eq. [6c] we set $\kappa_3$ (yellow) to the relaxed period of relatively high transmission, roughly from Memorial Day weekend through June 25, 2020. Eq. [6b] forms $\kappa_2$ (orange) as the average between these two levels of reduction. The most relaxed stage of transmission reduction we consider (blue), uses the same increment between the red and orange stages and between orange and yellow in Eq. [6d]. We do not separately estimate cocooning, but assume it matches the strictest level of reduction in the low-risk population when in the red stage, that observed during the stay-home period after March 24th for the orange and yellow stages, and then in the blue stage relaxes by a 25% increment, as shown in Eq. [6d], so that a reduction of 0.75 would drop to 0.6875.

**Algorithm 1:** Pseudo-code for the sampling procedure

Initialize $\Omega = \emptyset$
**while** $|\Omega| < 300$ **do**
  **for** $j = 1, \ldots, 6$ **do**
    **if** $j == 1$ **then**
      Generate a scenario (sample path) $\omega$ over time block $\mathcal{T}_1$
    **else**
      Continue simulating scenario $\omega$ over time block $\mathcal{T}_j$
    Compute $R^2 = 1 - \frac{\sum_{t \in \mathcal{T}_1 \cup \cdots \cup \mathcal{T}_j} (IH_t - \widehat{IH}_t)^2}{\sum_{t \in \mathcal{T}_1 \cup \cdots \cup \mathcal{T}_j} (IH_t - \overline{IH})^2}$
    **if** $R^2 < 0.85$ **then**
      break and initiate next $\omega$ and $j = 1$
  $\Omega \leftarrow \Omega \cup \{\omega\}$

### C Selection of Scenarios

After fitting the parameters via least-squares minimization using the procedure just described, we simulate a set of scenarios, $\Omega$. There are macro stochastics, involving modeling $\sigma$, $\omega_A$, $\gamma_A$, and $\gamma_Y$ as random variables; see Table 10. And, micro stochastics govern a binomially distributed number of transitions between compartments. Inevitably, some stochastic sample paths will yields hospitalizations that diverge from observed data, including sample paths in which spread quickly terminates after initializing with a single infectious person.

Here we describe how we select scenarios, sampled from the marginal distributions of $\sigma$, $\omega_A$, $\gamma_A$, and $\gamma_Y$, and from the micro-stochastics process, in order to generate a set of 300 scenarios, indexed by $\omega \in \Omega$ and used in Eqs. [1]. Because the projections at earlier time points guide the trigger optimization model at later time points, the goal is to start the latter with scenarios, $\omega \in \Omega$, that are consistent with observed hospitalizations up to that point in time. In order to evaluate the quality of scenario $\omega$ in this sense, we use

$$R^2 = 1 - \frac{\sum_t (IH_t - \widehat{IH}_t)^2}{\sum_t (IH_t - \overline{IH})^2},$$

where $\overline{IH}$ is the mean of the $IH_t$ values. Algorithm 1 summarizes the sampling procedure to generate $|\Omega| = 300$ scenarios.

Algorithm 1 uses the six time blocks $\mathcal{T}_j$, $j = 1, 2, 3, 4, 5, 6$, given in Table 3. We start with simulating a sample path $\omega$ during the first time block $\mathcal{T}_1$ (see line 4–5), and compute the corresponding $R^2$ value at the end of $\mathcal{T}_1$. If the scenario, $\omega$, has the desired quality over $\mathcal{T}_1$, i.e., $R^2 \geq 0.85$, we continue simulating the path $\omega$. At the end of each time block, we compute the corresponding $R^2$ value as shown in line 8, and decide whether the path is kept or discarded, as shown in lines 9–10. Here, the threshold value 0.85 of $R^2$ is a tuning parameter, which we return to below. Once Algorithm 1 provides 300 scenarios at the end of time block $\mathcal{T}_6$, we continue simulating them for use in the stochastic optimization model [3] over the rest of the time horizon.

Fig. 2 show histograms of the resulting marginal distributions from Algorithm 1 for $\sigma$, $\omega_A$, $\gamma_A$, and $\gamma_Y$. The scatter plots of Fig. 3 again use the scenarios, $\Omega$, obtained by Algorithm 1 to give insight to bivariate dependencies. We generate random variables from the independent nominal triangular distributions given in Table 10, and Algorithm 1 "accepts" those that are consistent with observed hospitalizations. The resulting marginal distributions can differ from the nominal distribution; see the distribution of $\omega_A$ in Fig. 2. In addition, even though we sample independently from the nominal distributions, Algorithm 1 can induce dependence; see the strong negative correlation between $\omega_A$ and $1/\gamma_Y$ in Fig. 3.

In Algorithm 1, we use a threshold of 0.85 for $R^2$, through each time block, to decide whether the generated path satisfies the desired quality through October 7, 2020. Of course, different thresholds are possible. To understand how sensitive the optimized policies are for different threshold values, we also obtain optimized policies using the scenarios generated with two more thresholds for $R^2$, 0.65 and 0.75, for Austin. Fig. 4 illustrates the projections of new daily hospitalizations and ICU hospitalizations. Analogs of the projections for those thresholds are also visualized in Fig. 1 in the main text for a threshold of 0.85. There are not dramatic differences across these three thresholds for $R^2$. The optimized policies are the same for thresholds of 0.65 and 0.75, and we observe a slightly smaller trigger (120 versus 130) for the red stage for an $R^2$ threshold of 0.85. As the threshold values shrink from 0.85 to 0.75 to 0.65: the expected number of days in the most stringent red-stage lock-down are 7.6, 5.4, and 5.8; the 95th percentiles of peak hospitalizations are 993, 1033, and 1028 (capacity is 1500); and the 95th percentiles of peak ICU demand are 317, 327, and 347 (capacity is 331). Here, the trigger policy is optimized for ICU capacity, and we observe occasional non-monotonic behavior due to statistical noise. That said, these results, along with Fig. 4, suggest that our results are not overly sensitive to our choice of 0.85 for the threshold.

For the purpose of visualizing results we use spaghetti plots that detail 300 sample paths, and we also use a single "representative" path. A simple least-squares selection of a path from the collection of 300 paths arguably does *not* yield a representative path. Because the timing of peaks in paths differs, the simple least-squares path is "flatter" through the cloud than most paths. Here we describe how we select a representative path from the collection of 300.

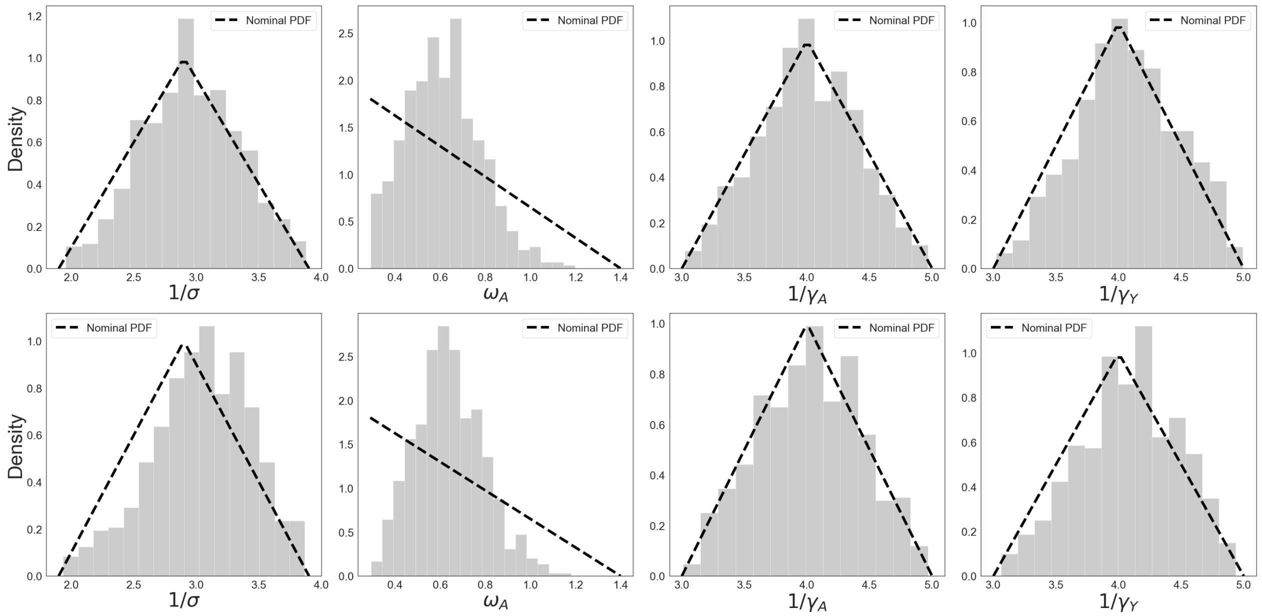


Supplementary Fig. 2: Histogram of the random variables $\sigma$, $\omega_A$, $\gamma_A$, and $\gamma_Y$ for the scenario set $\Omega$ obtained by Algorithm 1. The first and second row represent the set of scenarios for Austin and Houston, respectively. Rather than showing the rates, we show $1/\sigma$, $1/\gamma_A$, and $1/\gamma_Y$, which correspond to durations in days. We sample from the marginal distributions of each of the four parameters, and Algorithm 1 accepts sample paths consistent with hospitalizations, while also sampling a binomial number of transitions between compartments in the SEIR-style model. While there are modest deviations from the nominal triangular distributions for durations ($1/\sigma$, $1/\gamma_A$, and $1/\gamma_Y$), the distribution of the relative infectiousness of an asymptomatic individual ($\omega_A$) deviates significantly from its nominal distribution.

We define a selection criterion based on metrics of interest, such as the total number of people hospitalized in the general ward over the duration, the peak number of people in the hospital, and the time of that peak. We let $\widehat{IH}_{t,\omega}$, $\widehat{ICU}_{t,\omega}$, and $\widehat{H}_{t,\omega}$ denote the estimates of $IH_t$, $ICU_t$, and $H_t$ for each sample path, $\omega \in \Omega$, and we first obtain the squared deviations from the observed data, defining $Z_{\omega,obs}$ as follows:

$$Z_{\omega,obs} = \sum_t (\widehat{IH}_{t,\omega} - IH_t)^2 + w_{ICU}^2 \sum_t (\widehat{ICU}_{t,\omega} - ICU_t)^2 + w_H^2 \sum_t (\widehat{H}_{t,\omega} - H_t)^2,$$

where $w_{IH}$ and $w_{ICU}$ are scaling constants. Here, $t$ ranges over all days up to October 7, 2020, for which we have observed data. We then define metrics to represent the statistical properties of the scenarios as in Table 6.

| Metric | Definition | Standardized Metric |
|---|---|---|
| $IH_{\omega,tot} = \sum_{t=1}^{T} IH_{t,\omega}$ | Total number of patient-days | $Z_{\omega,IH_{tot}} = (IH_{\omega,tot} - \hat{\mu}_{IH_{tot}})/\hat{\sigma}_{IH_{tot}}$ |
| $IH_{\omega,max} = \max_t IH_{t,\omega}$ | Peak hospitalization | $Z_{\omega,IH_{max}} = (IH_{\omega,max} - \hat{\mu}_{IH_{max}})/\hat{\sigma}_{IH_{max}}$ |
| $IH_{\omega,med} = \text{median}_t IH_{t,\omega}$ | Median hospitalization | $Z_{\omega,IH_{med}} = (IH_{\omega,med} - \hat{\mu}_{IH_{med}})/\hat{\sigma}_{IH_{med}}$ |
| $t_{\omega,IH_{max}} = \arg\max_t IH_{t,\omega}$ | Timing of hospitalization peak | $Z_{t_{\omega,IH_{max}}} = (t_{\omega,IH_{max}} - \hat{\mu}_{t_{IH_{max}}})/\hat{\sigma}_{t_{IH_{max}}}$ |

Supplementary Table 6: Metrics for hospitalized patients in the general ward under each sample path $\omega$. Note that $\hat{\mu}$ and $\hat{\sigma}$ represent the sample mean and sample standard deviation for each metric, across the $|\Omega| = 300$ sample paths.

Analogs of the standardized metrics for those hospitalized in the general ward ($IH$) from Table 6 are also computed for those hospitalized in the ICU ($ICU$), and new daily hospital admissions ($H$) to form a total of 12 such metrics.

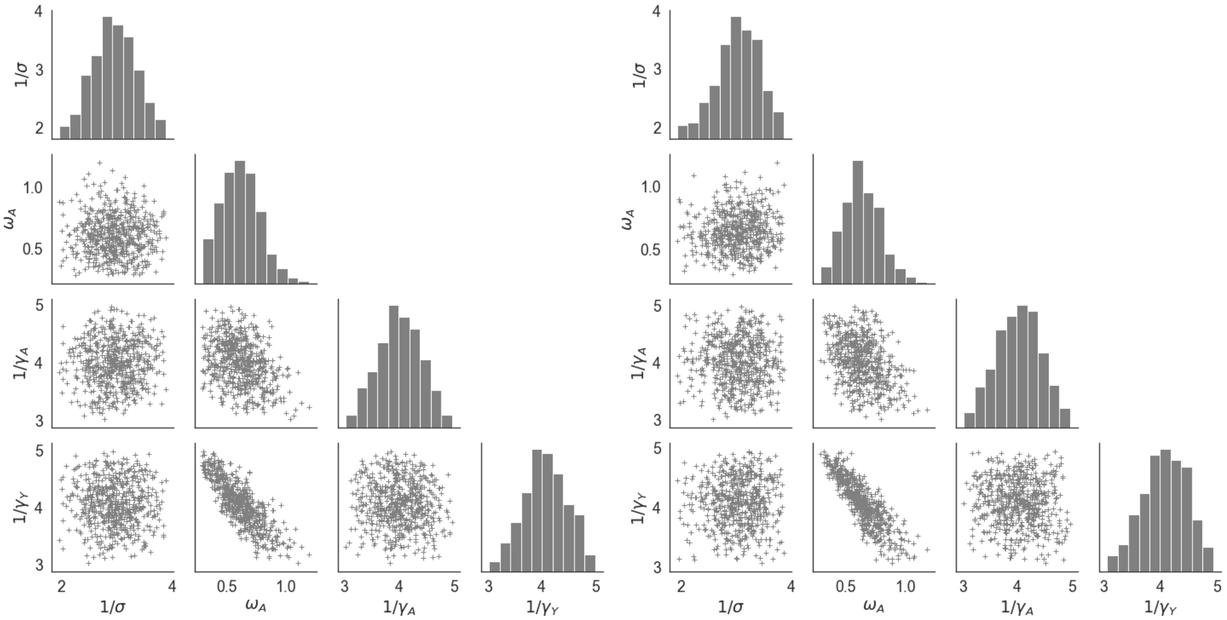


Supplementary Fig. 3: Scatter plot of the random variables $1/\sigma$, $\omega_A$, $1/\gamma_A$, and $1/\gamma_Y$ for the scenario set $\Omega$ selected by Algorithm 1. The left and right scatter plots represent the set of scenarios, $\Omega$, for Austin and Houston, respectively. We sample *independently* from the marginal distributions of each of the four parameters, and the algorithm accepts sample paths consistent with hospitalizations. This can induce dependencies as shown, for example, between $1/\gamma_Y$ and $\omega_A$.

We select the representative scenario $\omega'$ such that

$$\omega' \in \underset{\omega \in \Omega}{\arg\min} \left( \frac{w_{std}}{12} Z_{\omega,std} + w_{obs} Z_{\omega,obs} \right) \quad [7]$$

where

$$\begin{aligned} Z_{\omega,std} = {} & Z^2_{\omega,IH_{tot}} + Z^2_{\omega,IH_{max}} + Z^2_{\omega,IH_{med}} + Z^2_{\omega,t_{IH_{max}}} \\ & + Z^2_{\omega,ICU_{tot}} + Z^2_{\omega,ICU_{max}} + Z^2_{\omega,ICU_{med}} + Z^2_{\omega,t_{ICU_{max}}} \\ & + Z^2_{\omega,H_{tot}} + Z^2_{\omega,H_{max}} + Z^2_{\omega,H_{med}} + Z^2_{\omega,t_{H_{max}}}, \end{aligned}$$

where $w_{std}$ and $w_{obs}$ are positive weights that sum to one. This method is used to select the representative paths plotted in Fig. 1 in the main text, and Figs. 4-5 and Figs. 8-9. Moreover, when we have paired plots of total hospitalizations, or ICU hospitalizations, along with daily admissions (see Fig. 1 in the main text and Figs. 4-5) then the cyan and black curves correspond to the same scenario.

### D Test Settings

#### D.1 Benchmark Policies

In Section 2 of the main text we compare four alternative policies to our optimized four-stage trigger policy. First, we test a benchmark which is again optimized and differs from the four-stage policy only in that it has access to just two stages of mitigation, red and yellow. In this case, we require a single threshold to identify when to toggle between the two stages. In our second benchmark, we again optimize in the same way as the four-stage policy, but we do so using a 0.95-level probabilistic constraint to respect *total* hospital capacity rather than ICU capacity.

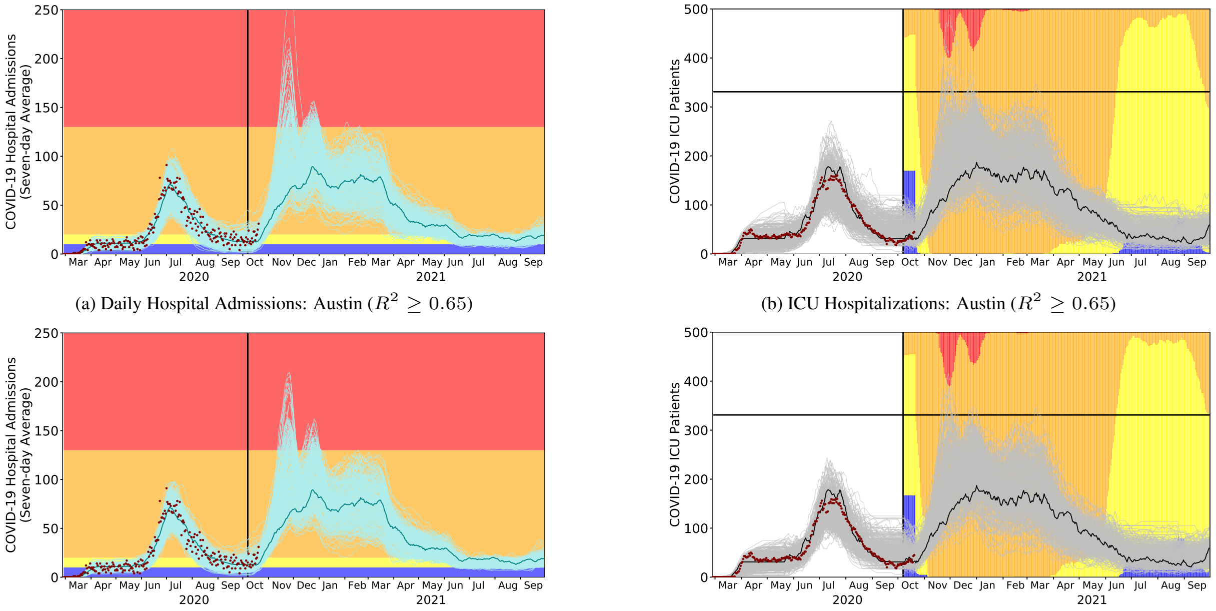


(a) Daily Hospital Admissions: Austin ($R^2 \geq 0.65$)

(b) ICU Hospitalizations: Austin ($R^2 \geq 0.65$)

(c) Daily Hospital Admissions: Austin ($R^2 \geq 0.75$)

(d) ICU Hospitalizations: Austin ($R^2 \geq 0.75$)

Supplementary Fig. 4: COVID-19 healthcare projections for Austin under the optimized staging policy, from October 7, 2020 through September 30, 2021. Each row represents a projection with a different threshold value for $R^2$ in Algorithm 1, and are analogs of Fig. 1 in the main text, which uses $R^2 \geq 0.85$. The strategies were derived to minimize the expected days in costly alert stages while respecting ICU capacity. The light curves indicate 300 stochastic simulations, the single solid curves are representative central projections, the red points correspond to the reported COVID-19 admissions and ICU census for all Austin area hospitals through October 20, 2020, and the vertical black line indicates the start of the projection period. (a,c) The optimized policy triggers changes in the COVID-19 alert level when the seven-day moving average of daily COVID-19 hospital admissions crosses optimized thresholds, as indicated by the colored horizontal bands. The alert stage shifts when the admissions indicator surpasses or recedes below the corresponding threshold. (b,d) The policy provides a 95% guarantee that the number of COVID-19 ICU patients does not exceed the estimated local capacity of 331 patients (black horizontal line). The background colors represent the proportion of the simulated scenarios in each alert stage on each day. Optimized policies that keep us within ICU capacity are identical under thresholds 0.65 and 0.75, and there is a modest change in the red stage's trigger (represented by the red horizontal band) for a threshold of 0.85.

We assess two additional benchmarks. Instead of triggering based on the seven-day moving average of daily hospital admissions, we use the seven-day moving average of (i) daily census in the ICU and (ii) the estimated prevalence of infectious individuals. Alternative (i) is currently used in France [13], and in this case we use respective trigger thresholds for the red, orange, and yellow stages based on 60%, 30%, and 0% of the ICU capacity. Alternative (ii) was proposed by Harvard Global Health Institute [14], and stages red, orange, yellow, and blue correspond to more than 25, 10 to 25, 1 to 10, and less than 1 daily confirmed infectious case(s) per 100 000 people. Those thresholds are adjusted with the population of Austin MSA (2.2 million) and Houston MSA (7.1 million), and use a 1-in-10 ratio between the confirmed infectious cases and total estimated infectious cases.

#### D.2 ACS Parameters

Table 7 details parameters used in the ACS analysis in Section E.2 for the Austin MSA, involving weighted combinations of transmission reduction between the orange and yellow stages. The stages differ in the physical distancing coefficient, $\kappa$, but all use the same cocooning coefficient, $c$, as shown in the table.

We assume the ACS can be set up within two days; i.e., the ACS will be ready to use two days after the trigger threshold is reached. In reality, an ACS is built in phases with staffing being the final phase that allows the facility to function. The trigger threshold to operationalize and size the ACS are both selected by a grid search with the grid step

| Transmission Reduction | | $\kappa$ | $c$ |
|---|---|---|---|
| orange | yellow | | |
| 75% | 25% | 0.6697 | 0.7436 |
| 50% | 50% | 0.6473 | 0.7436 |
| 25% | 75% | 0.6250 | 0.7436 |
| 0% | 100% | 0.6026 | 0.7436 |

Supplementary Table 7: The two left-most columns indicate the relative weight on transmission reduction factors for the orange and yellow stages. So, the last row corresponds to yellow, while the 50-50% row corresponds to equal weight on the orange and yellow values. The two right-most columns show the resulting transmission reduction coefficients for the low-risk population ($\kappa$) and the high-risk population ($c$) for Austin.

sizes of 10 and 100, respectively

### E Supplementary Analysis

#### E.1 Optimal Triggers: Houston

We present results for Houston in a parallel format as for Austin in Section 2 of the main text. Houston's nine-county MSA has a population of 7.1 million. We use the estimated transmission rates from February 19–October 7, 2020 as given in Tables 3 and 5. Detailed results for our proposed policy and the four benchmarks are shown in Table 8 and we visualize the disease dynamics associated with the optimal triggers by new daily hospitalizations and the ICU daily census in Fig. 5, parallel to Table 1 and Fig. 1 in the main text. We also visualize the benchmark comparison results in Fig. 6, like in Fig. 2 in the main text. Qualitatively the results are similar to those for Austin except that for Houston the *Percent ICU* policy is reliable, albeit at a larger socioeconomic cost than the *Optimal* policy.

| | | Policies | | | | |
|---|---|---|---|---|---|---|
| | | Optimal (ICU capacity) | Optimal two-stage (ICU capacity) | Optimal hospital (overall capacity) | Percent ICU (France) | Incidence (Harvard) |
| Indicator data | | COVID-19 hospital admissions (seven-day average) | | | percent ICU beds occupied by COVID-19 | new cases per 100 000 (seven-day average) |
| Thresholds | blue (new normal) | <30 | — | <30 | — | <1 |
| | yellow (moderate risk) | 30-90 | <380 | 30-160 | <30% | 1-10 |
| | orange (high risk) | 90-500 | — | 160-560 | 30%-60% | 10-25 |
| | red (very high risk) | >500 | >380 | >560 | >60% | >25 |
| Median days in red stage [90% PI] | | 0 [0-0] | 15 [14-30] | 0 [0-0] | 16 [0-23] | 0 [0-14] |
| Probability ICU demand exceeds capacity | | 2.3% | 2.3% | 53.7% | 0.0% | 0.0% |
| Median peak ICU demand [patients] | | 780 | 901 | 1010 | 658 | 303 |
| 95th percentile of peak ICU demand | | 977 | 984 | 1187 | 740 | 395 |
| Median unserved ICU demand (patient-days) [90% PI] | | 0 [0-0] | 0 [0-0] | 15 [0-3915] | 0 [0-0] | 0 [0-0] |
| 99th percentile of unserved ICU demand (patient-days) | | 540 | 24 | 6843 | 0 | 0 |

Supplementary Table 8: Performance comparison across five COVID-19 staging policies for Houston. From left to right, the three optimal policies are derived to prevent overwhelming ICU demand using either a four-level or two-level alert system or to prevent overwhelming inpatient demand using a four-level system. As benchmarks, we evaluate policies implemented in France [13] and proposed as gating criteria for relaxing measures and opening schools in [14]. For Houston, the orange and red thresholds for the *Percent ICU* policy translate to 300 and 600 COVID-19 ICU cases, respectively; the yellow, orange, and red thresholds for the *Incidence* policy translate to 710, 7100, and 17750 new cases, respectively, assuming that one in ten cases is reported. We implemented each policy in our stochastic SEIR model fit to hospitalization data for the Houston, Texas MSA assuming the reported COVID-19 ICU and inpatient capacities of 1000 and 4500 beds, respectively. Outcomes are based on 300 stochastic simulations of COVID-19 transmission and healthcare burden from October 7, 2020 through September 30, 2021 under each policy.

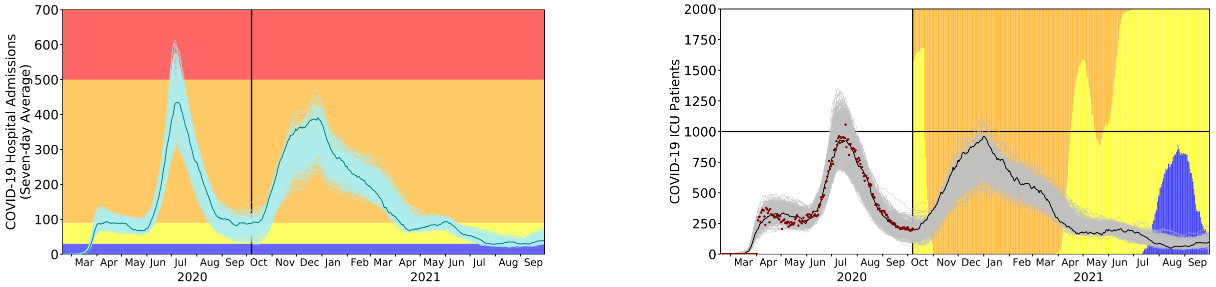


(a) Daily Hospital Admissions: Houston

(b) ICU Hospitalizations: Houston

Supplementary Fig. 5: COVID-19 healthcare projections for Houston under the optimized staging policy, from October 7, 2020 (marked by the vertical line) through September 30, 2021. The trigger strategy was derived to minimize the expected days in costly alert stages while respecting ICU capacity. In both plots, the light curves indicate 300 stochastic simulations, the single solid curve is a representative *central* projection, and the red points correspond to the ICU census for all Houston MSA hospitals through October 20, 2020. (a) The optimized policy triggers changes in the COVID-19 alert level when the seven-day moving average of daily COVID-19 hospital admissions crosses set thresholds, as indicated by the colored horizontal bands. The alert stage shifts when the admissions indicator surpasses or recedes below the corresponding threshold. (b) The policy provides a 95% guarantee that the number of COVID-19 ICU patients does not exceed the estimated local capacity of 1000 patients (black horizontal line). The background colors represent the proportion of the simulated scenarios in each alert stage on each day.

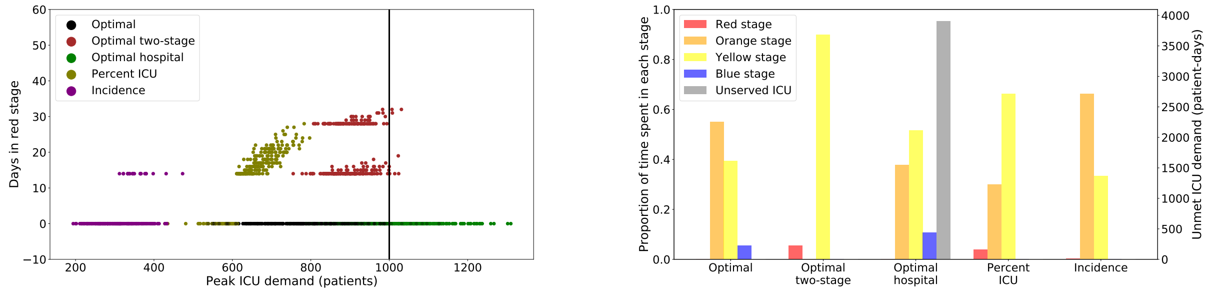


(a) Days in lock-down (red) versus peak ICU demand

(b) Proportion of time per stage and unserved ICU patient-days (95th percentile)

Supplementary Fig. 6: Projected ICU surges and days under lock-down for the optimized strategy versus four alternative policies. *Optimal* is the recommended strategy. *Optimal two-stage* is optimized to respect ICU capacity under a two-level alert system, and *Optimal hospital* respects total hospital capacity under a four-level alert system. *Percent ICU* is based on France's lock-down policy [13] and *Incidence* is based on reopening criteria proposed by Harvard [14]. (a) The maximum daily number of COVID-19 patients in ICUs versus the number of days under the most restrictive alert level of red. Each point represents the result of a single stochastic simulation under one of the five policies (indicated by color). The plot includes 300 points per policy; the vertical gray line indicates the estimated COVID-19 ICU capacity of 1000 patients for the Houston area. The *Optimal* policy is designed to minimize the use of the strictest stages of mitigation while having 95% of the peak-demand values within ICU capacity. (b) The expected proportion of days spent in each stage, colored in the same manner as Fig. 5 and the 95th percentile of ICU shortage measured in patient-days above capacity, in gray with values indicated on the right $y$-axis.

#### E.2 Construction of an Alternate Care Site

The success of trigger-based policies for ICU and hospital capacity depends on public adherence, which is not guaranteed. In case of failure, Austin applied this framework to determine both a threshold for launching an alternate care site (ACS) and the required capacity. The city planned to transfer low acuity COVID-19 patients to the ACS when cases exceed the city-wide estimated 1500 patient capacity. We consider four scenarios for non-adherence, each assuming a constant transmission rate between those estimated for Austin under restrictions of the orange and yellow stages. The lowest adherence scenario assumes the transmission rate estimated in Austin from Memorial Day (late-May) through late-June of 2020, after *Opening Up America Again* was promulgated and before the city and state took measures to curb the early summer surges, including strict face-mask requirements. In this situation, the ACS should be stood up when the seven-day rolling average of COVID-19 hospital admissions reaches 110; the recommended trigger increases to 220 under the highest adherence scenario (Table 9 and Fig. 7). Failing to construct an ACS would lead to a median of 30519 [90% PI: 11834-62450] and 0.0 [90% PI: 0-255] patient-days of unmet demand, under these two scenarios, respectively. We would expect demand for ACS beds to last up to two months, and to decrease with higher levels of adherence.

| Scenario | | Launch | Target | Probability | 95th Percentile | |
|---|---|---|---|---|---|---|
| Adherence | Transmission rate | trigger | capacity | launched | Days used | Peak demand |
| moderate-high | 75%-25% | 220 | 100 | 0.09 | 41 | 55 |
| moderate | 50%-50% | 200 | 700 | 0.64 | 61 | 640 |
| moderate-low | 25%-75% | 210 | 1400 | 0.97 | 63 | 1292 |
| low | 0%-100% | 110 | 2000 | 1.00 | 65 | 1906 |

Supplementary Table 9: Optimized ACS policies under four adherence scenarios for the Austin area. The scenarios assume that the public do not fully comply with enacted alert levels, resulting in a constant transmission rate lying somewhere between those expected under the orange and yellow stages. The percents in the second column indicate the relative weighting of the orange and yellow rates; for example, the moderate-high rate is obtained via a 75-25 weighting of the two rates, respectively. The ACS policies track the moving seven-day average of COVID-19 hospital admissions and assume that the ACS will be ready to accept patients two days after triggering. The *launch trigger* is the derived threshold for opening the ACS; the *target capacity* is the estimated total ACS beds needed, based on the criteria that 95% of projections remain under ACS capacity; *probability launched* is the fraction of simulations in which admissions exceed the launch trigger; the 95th percentiles of *days used* and *peak demand* provide upper bounds on the projected demand. Coupled optimization of the the trigger and capacity in discrete steps of 100 beds and 10 daily admissions, respectively, along with the resulting slack between peak demand and target capacity can produce small non-monotonicities in optimal triggers, as seen in the middle two rows. We assume the ACS remains open for 100 days once triggered, which is more than sufficient to cover the estimated 95th percentile of days required (penultimate column).

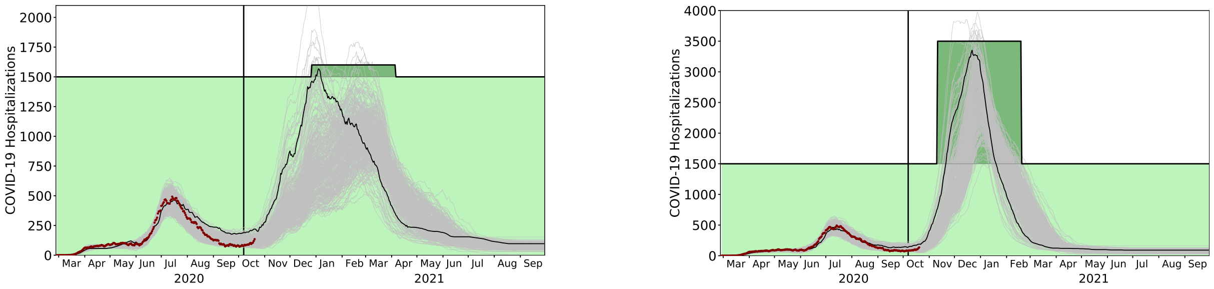


(a)

(b)

Supplementary Fig. 7: Projected COVID-19 hospital census for Austin under optimized ACS policies, from October 7, 2020 through September 30, 2021. In both plots, the light curves indicate 300 stochastic simulations, the single solid curve is a relatively *early* projection, light green indicates the baseline COVID-19 hospital capacity, dark green indicates the timing and size of the ACS triggered by the solid curve, the red points correspond to the reported COVID-19 hospital census for all Austin area hospitals through October 20, 2020, and the vertical black line indicates the start of the projection period. (a) Under a moderate-high adherence scenario, a 100-bed ACS is launched when the seven-day rolling average of hospital admissions exceeds 220. (b) Under a low adherence scenario, a 2000-bed ACS is launched when the seven-day rolling average of hospital admissions exceeds 110. Hospital admissions are not shown.

#### E.3 Sensitivity Analysis

We report additional results on the sensitivity associated with public compliance with recommended physical distancing, or lack thereof. Figs. 8(a) and 8(b) show results assuming that after October 7th the transmission reduction level stays at that of the yellow stage. The ICU hospitalizations greatly exceed the ICU capacity in all 300 scenarios for both Austin and Houston. Median peak ICU hospitalizations are 810 for Austin (capacity 331) and 2574 for Houston (capacity 1000) with corresponding 95th percentiles of 1094 and 2920. For Austin and Houston we estimate a median 20366 and 70274 unserved patient-days in the ICU, with respective 90% prediction intervals of [13505-30575] and [55077-84364].

Figs. 8(c) and 8(d) show results that suppose optimized triggers from Table 1 in the main text are carried out but the high-risk population is not cocooned, and is instead subject to transmission rates in the low-risk population. In this case, we violate ICU capacity in 4.3% of the 300 out-of-sample scenarios in Austin and 97.3% of the scenarios in Houston. The 95th percentile of peak ICU hospitalizations grows to 329 for Austin (capacity 331) and 1231 for Houston (capacity 1000). After October 7, 2020, for Austin, we estimate a median 0 unserved patient-days, and for Houston, the corresponding estimate is 1626, with respective 90% prediction intervals of [0-0] and [76-4014].

In Fig. 9 we show results of tests for four situations in which the public complies with the optimal policy up to a certain date and after that date, compliance fails and transmission matches that of the yellow stage. The figure shows results when such non-compliance respectively begins on February 1, March 1, April 1, and May 1, 2021.

For Austin starting from February 1st, March 1st, and April 1st, and Houston starting from February 1st, the surge results in the 95th percentile of the number of ICU patients exceeding capacity. Detailed simulation results, which include the number of unserved patient-days and deaths follow. We compute 90% prediction intervals using symmetric order statistics, i.e., the middle 270 of 300 scenarios. As a result, the interval for unserved ICU patient-days is necessarily [0-0] whenever the 95th percentile of peak ICU demand is within capacity.

**Austin:**
Yellow stage starts on:

- February 1, 2021:
  - probability the number of hospitalized ICU patients exceeds COVID-19 ICU capacity: 88.7%
  - 95th percentile of peak ICU hospitalizations: 555
  - median unserved ICU patient-days: 2236
  - 90% prediction interval of unserved ICU patient-days: [0-7368]
- March 1, 2021:
  - probability the number of hospitalized ICU patients exceeds COVID-19 ICU capacity: 53.7%
  - 95th percentile of peak ICU hospitalizations: 431
  - median unserved ICU patient-days: 8
  - 90% prediction interval of unserved ICU patient-days: [0-2368]
- April 1, 2021:
  - probability the number of hospitalized ICU patients exceeds COVID-19 capacity: 5.3%
  - 95th percentile of peak ICU hospitalizations: 333
  - median unserved ICU patient-days: 0
  - 90% prediction interval of unserved ICU patient-days: [0-2]
- May 1, 2021:
  - probability the number of hospitalized patients exceeds COVID-19 capacity: 2.7%
  - 95th percentile of peak ICU hospitalizations: 317
  - median unserved ICU patient-days: 0

    - 90% prediction interval of unserved ICU patient-days: [0-0]

**Houston:**
Yellow stage starts on:

- February 1, 2021:
    - probability the number of hospitalized ICU patients exceeds COVID-19 capacity: 74.0%
    - 95th percentile of peak ICU hospitalizations: 1294
    - median unserved ICU patient-days: 1045
    - 90% prediction interval of unserved ICU patient-days: [0-8941]
- March 1, 2021:
    - probability the number of hospitalized ICU patients exceeds COVID-19 capacity: 2.7%
    - 95th percentile of peak ICU hospitalizations: 978
    - median unserved ICU patient-days: 0
    - 90% prediction interval of unserved ICU patient-days: [0-0]
- April 1, 2021:
    - probability the number of hospitalized ICU patients exceeds COVID-19 capacity: 2.3%
    - 95th percentile of peak ICU hospitalizations: 977
    - median unserved ICU patient-days: 0
    - 90% prediction interval of unserved ICU patient-days: [0-0]
- May 1, 2021:
    - probability the number of hospitalized ICU patients exceeds COVID-19 capacity: 2.3%
    - 95th percentile of peak ICU hospitalizations: 977
    - median unserved ICU patient-days: 0
    - 90% prediction interval of unserved ICU patient-days: [0-0]

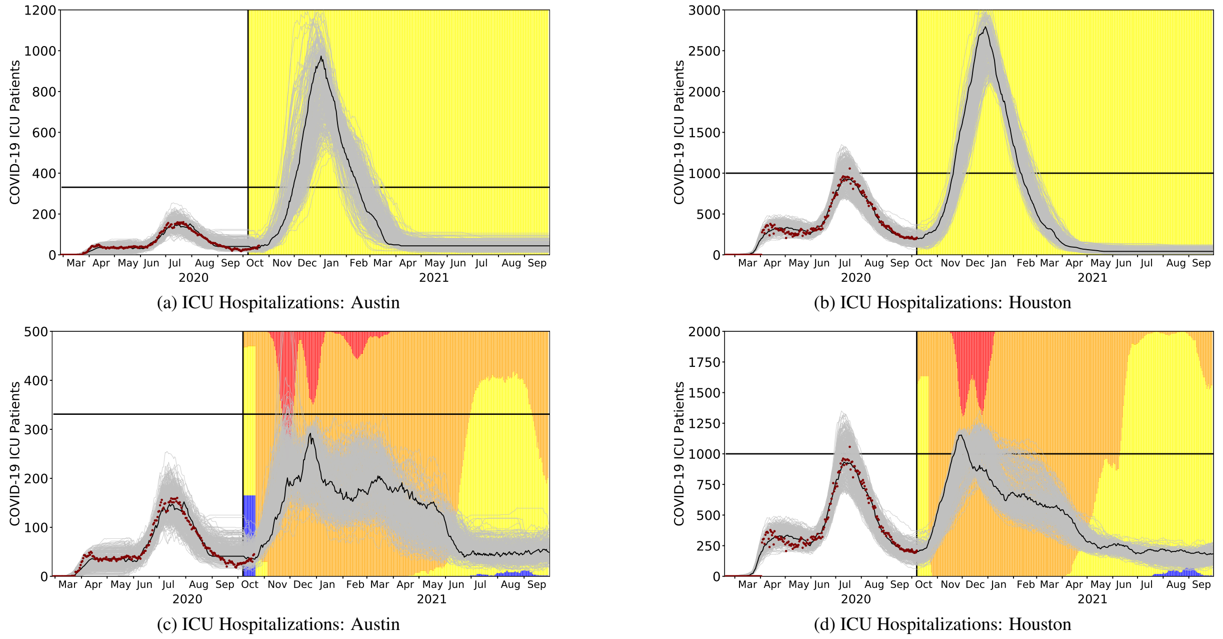


(a) ICU Hospitalizations: Austin

(b) ICU Hospitalizations: Houston

(c) ICU Hospitalizations: Austin

(d) ICU Hospitalizations: Houston

Supplementary Fig. 8: Projections for COVID-19 ICU hospitalizations in the Austin MSA and Houston MSA when public compliance wanes, and high-risk population cocooning is not followed through the time horizon. Panels (a) and (b) present daily ICU hospitalizations (heads in beds) when the yellow stage holds after October 7th, and panels (c) and (d) present daily ICU hospitalizations when optimized triggers are used but when there is no cocooning; instead transmission reduction in the high-risk population degenerates to that of the low-risk population.

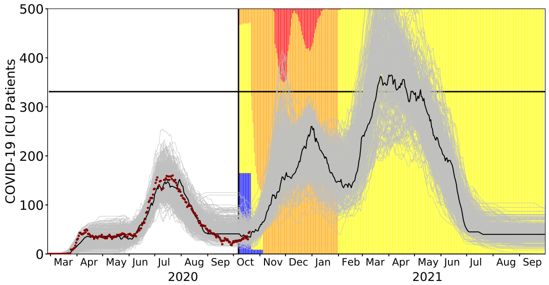


(a) Compliance until February 1st: Austin

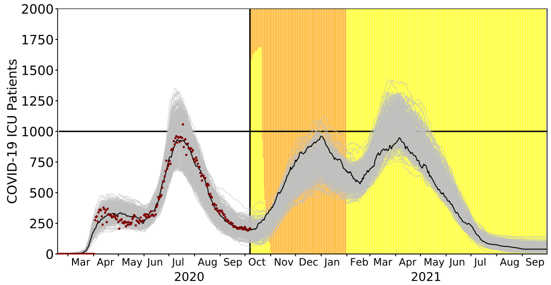


(b) Compliance until February 1st: Houston

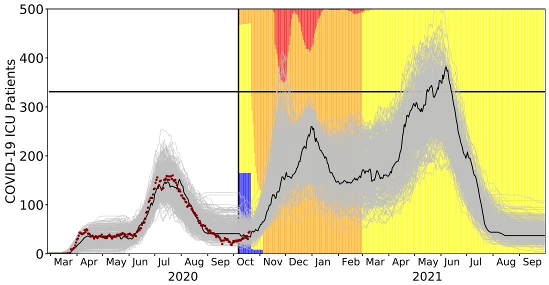


(c) Compliance until March 1st: Austin

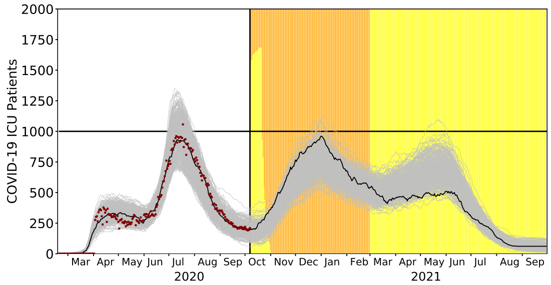


(d) Compliance until March 1st: Houston

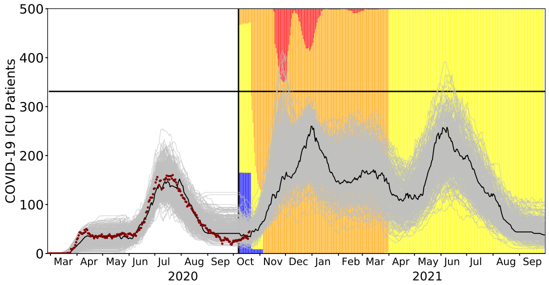


(e) Compliance until April 1st: Austin

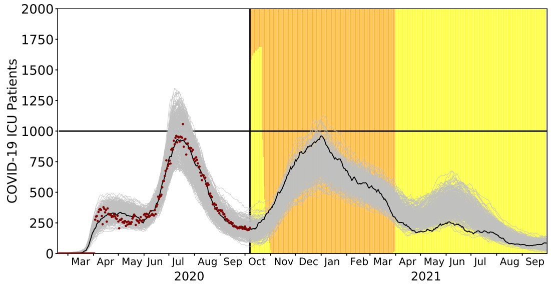


(f) Compliance until April 1st: Houston

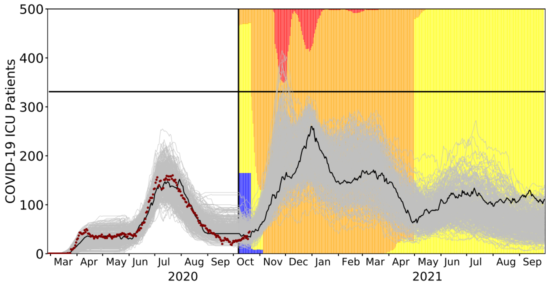


(g) Compliance until May 1st: Austin

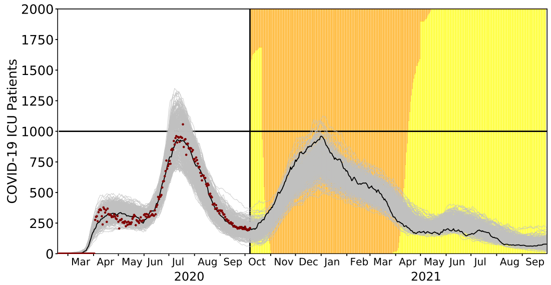


(h) Compliance until May 1st: Houston

Supplementary Fig. 9: Projections for daily COVID-19 ICU hospitalizations (heads in beds) in the Austin MSA and Houston MSA when the optimal policy in Table 1 of the main text is in effect until: February 1st (a) and (b); March 1st (c) and (d); April 1st (e) and (f); and May 1st (g) and (h). The level of transmission corresponds to the yellow stage thereafter. The projections suggest that, to stay within total hospital capacity, public compliance is needed through May 1st in Austin and March 1st in Houston.

| Parameters | Values | Source |
|---|---|---|
| $\beta$: transmission rate | Austin: 0.06901<br>Houston: 0.06401 | [4, 15] |
| $P$: proportion of pre-symptomatic transmission (%) | 44 | [5] |
| $\omega_A$: infectiousness of individuals in compartment $IA$, relative to $IY$ | $\omega_A \sim$ Triangular (0.29,0.29,1.4) | [16] |
| $\tau$ : symptomatic proportion (%) | 57 | [17] |
| $\omega_P$: infectiousness of individuals in pre-symptomatic and pre-asymptomatic compartments, relative to symptomatic and asymptomatic compartments | $\omega_P = \frac{P}{1-P} \frac{\tau(\frac{YHR}{\eta_H}+\frac{1-YHR}{\gamma_Y})+(1-\tau)\frac{\omega_A}{\gamma_A}}{\frac{\tau}{\rho_Y}+(1-\tau)\frac{\omega_A}{\rho_A}}$ | |
| $\sigma$ : exposed rate | $\frac{1}{\sigma} \sim$ Triangular (1.9, 2.9, 3.9) | Based on incubation [18] and pre-symptomatic periods |
| $\gamma_A$: recovery rate from compartment $IA$ | $\frac{1}{\gamma_A} \sim$ Triangular (3, 4, 5) | [5] |
| $\gamma_Y$: recovery rate from symptomatic compartment $IY$ | $\frac{1}{\gamma_Y} \sim$ Triangular (3, 4, 5) | [5] |
| $\rho_A$: rate at which pre-asymptomatic individuals become asymptomatic | Equal to $\rho_Y$ | [5] |
| $\rho_Y$: rate at which pre-symptomatic individuals become symptomatic | $\frac{1}{\rho_Y} = 2.3$ | [5] |
| $IFR$: infected fatality ratio, age specific (%) | Low risk / High risk:<br>0.000917 / 0.00917<br>0.00218 / 0.0218<br>0.0339 / 0.339<br>0.252 / 2.52<br>0.644 / 6.44 | Age adjusted from [19] |
| $YFR$: symptomatic fatality ratio, age specific (%) | Low risk / High risk:<br>0.00161 / 0.0161<br>0.00382 / 0.0382<br>0.0594 / 0.594<br>0.442 / 4.42<br>1.13 / 11.3 | $YFR = \frac{IFR}{1-\tau}$ |
| $C$: daily cost to stay at specific stages | $C_1$ 10000<br>$C_2$ 100<br>$C_3$ 10<br>$C_4$ 1 | |

Supplementary Table 10: Model parameters

| Parameters | Value | Source |
|---|---|---|
| $\eta_H$ : rate from symptom onset to hospital admission | 0.1695 | 5.9 day average from symptom onset to hospital admission [20] |
| $YHR$: symptomatic case hospitalization rate (%) | Low risk / High risk<br>0.0279 / 0.2791<br>0.0215 / 0.2146<br>1.3215 / 13.2514<br>2.8563 / 28.5634<br>3.3873 / 33.8730 | Age adjusted from [19] |
| $p_{IH}$ | Fitted time series, starting at 0.6717 | hospital system data |
| $\gamma_H, \gamma_{ICU}$ : recovery rate in compartment $IH$ and $ICU$ | Fitted parameters | hospital system data |
| $\pi$ : rate symptomatic individuals go to hospital, age-specific | $\pi = \frac{\gamma_Y \cdot YHR}{\eta_H + (\gamma_Y - \eta_H)YHR}$ | |
| $\eta_{ICU}$ : rate from hospital admission to ICU | A time series which is constant specific to time blocks | hospital system data |
| $\mu$ : rate from ICU to death | Fitted parameters | hospital system data |
| $ICUFR$: ICU death ratio, age specific (%) | $ICUFR$<br>5.8592<br>5.8592<br>5.8592<br>15.6207<br>30.8526 | hospital system data |
| $HICUR$: hospitalized ICU ratio | A time series with a decreasing rate specific to time blocks, starting at 0.1574 | hospital system data |
| $\nu_H$ : ICU rate on hospitalized individuals, age-specific | $\nu_H = \frac{\gamma_H * HICUR}{\eta_{ICU} + (\gamma_H - \eta_{ICU})HICUR}$ | |
| $\nu_{ICU}$ : death rate on ICU individuals, age-specifc | $\nu_{ICU} = \frac{\gamma_{ICU} * ICUFR}{\mu + (\gamma_{ICU} - \mu)ICUFR}$ | |
| $B$: Total hospital bed capacity (including ICU) | Austin: 1500<br>Houston: 4500 | Estimates provided by each of the region's hospital systems and aggregated by regional public health leaders |
| $B_{ICU}$: ICU capacity | Austin: 331<br>Houston: 1250 | Estimates provided by each of the region's hospital systems and aggregated by regional public health leaders |
| $1_{\{\text{school closure}\}}$: school closure dates | Austin: $3/19/2020 - 9/8/2020$, $5/26/2021 - 8/23/2021$<br>Houston: $3/19/2020 - 9/8/2020$, $5/28/2021 - 8/23/2021$ | |

Supplementary Table 11: Hospitalization parameters

### Acknowledgements

The use of the hospital system data to generate the input model parameters (in Appendix B) was approved as exempt human subjects research by the IRB of The University of Texas at Austin.
